# Rapid systematic review of the sensitivity of SARS-CoV-2 molecular testing on saliva compared to nasopharyngeal swabs

**DOI:** 10.1101/2020.08.05.20168716

**Authors:** E. Peeters, S. Kaur Dhillon Ajit Singh, J. Vandesompele, P. Mestdagh, V. Hutse, M. Arbyn

## Abstract

**Background:** Nasopharyngeal sampling has been the standard collection method for COVID-19 testing. Due to its invasive nature and risk of contamination for health care workers who collect the sample, non-invasive and safe sampling methods like saliva, can be used alternatively.

**Methods:** A rapid systematic search was performed in PubMed and medRxiv, with the last retrieval on June 6^th^, 2020. Studies were included if they compared saliva with nasopharyngeal sampling for the detection of SARS-CoV-2 RNA using the same RT-qPCR applied on both types of samples. The primary outcome of interest was the relative sensitivity of SARS-CoV-2 testing on saliva versus nasopharyngeal samples (used as the comparator test). A secondary outcome was the proportion of nasopharyngeal-positive patients that tested also positive on a saliva sample.

**Results:** Eight studies were included comprising 1070 saliva-nasopharyngeal sample pairs allowing assessment of the first outcome. The relative sensitivity of SARS-CoV-2 testing on saliva versus nasopharyngeal samples was 0.97 (95% CI=0.92-1.02). The second outcome incorporated patient data (n=257) from four other studies (n=97 patients) pooled with four studies from the first outcome (n=160 patients). This resulted in a pooled proportion of nasopharyngeal positive cases that was also positive on saliva of 86% (95% CI=77-93%).

**Discussion:** Saliva could potentially be considered as an alternative sampling method when compared to nasopharyngeal swabs. However, studies included in this review often were small and involved inclusion of subjects with insufficient information on clinical covariates. Most studies included patients who were symptomatic (78%, 911/1167). Therefore, additional and larger studies should be performed to verify the relative performance of saliva in the context of screening of asymptomatic populations and contact-tracing.

## Introduction

Adequate testing for tracking and tracing of COVID-19 cases, is an effective strategy to mitigate the current SARS-CoV-2 pandemic.^1^ Currently, collecting and testing samples from lower respiratory tract (i.e. sputum or endotracheal aspirate) or upper respiratory tract (i.e. nasopharyngeal (NP) swabs), are recommended for COVID-19 testing by the (European) Centers for Disease Control and Prevention and World Health Organization.^2,3^ The collected specimen, preferably stored in a liquid storage medium, is then sent to a laboratory for the detection of viral RNA using reverse transcription quantitative polymerase chain reaction (RT-qPCR). Before the SARS-CoV-2 outbreak, studies related to SARS-CoV and other respiratory viruses, have reported the potential use of saliva as a source for viral RNA testing.^4-6^ Biologically, this can be explained by the expression of angiotensin-converting enzyme 2 (ACE2) receptors to which SARS-CoV-2 binds and that are present in epithelial cells of the oral mucosa.^7,8^

The non-invasive nature of collecting saliva samples from suspected individuals provides an advantage when compared to NP sample collection. Collection of saliva samples requires individuals to either spit in a cup or swabbing the mucosa of the oral cavity or tongue. This offers more comfort, ease and is quicker when compared to NP swabs as the latter often leads to discomfort, pain and may sometimes lead to bleeding.^9^ Furthermore, saliva sampling methods may be less aerosol-generating and thereby reduces the risk of infection to health care workers responsible to take the sample.^10,11^ Additionally, patients can collect saliva samples themselves without the need of a physician or nurse.^12^ All these advantages make saliva collection more suitable for repeated testing and may overcome certain barriers inherent to NP sampling. To assess the diagnostic sensitivity of SARS-CoV-2 testing on saliva compared to NP swabs, we conducted a rapid systematic review.

## METHODS

### Study selection criteria

Studies were included if subjects were tested using a RT-qPCR method detecting RNA of the SARS-CoV-2 virus on saliva samples and NP samples. Covariates of interest were: study design, severity of symptoms, hospitalization status, and timing of testing (days after onset of symptoms). We only selected paired studies, where the two types of specimen were collected from the same patients.

### Systematic search

A rapid systematic search was performed using public bibliographic databases containing peer-reviewed and non-peer reviewed articles from PubMed and medRxiv, respectively. The reference lists from the included articles were also screened. In addition, we verified the reference lists of other relevant reviews.^10,12-19^

### Data analysis

Patient-paired data was used to construct 2×2 contingency tables (see template in Table 1). The primary outcome of interest was the relative virological sensitivity of SARS-CoV-2 testing on saliva vs NP swabs. This relative sensitivity was calculated by dividing the sensitivity of testing on saliva (SEN_saliva_+=[a+b]/[a+b+c]) by the sensitivity on NP swabs (SEN_NP_+=[a+c]/[a+b+c]), assessed in populations where both types of samples were collected at the same time. Given the paired design, the denominators of the two proportions are the same and cancel each other out. Therefore, the relative sensitivity SEN_saliva+_ / SEN_NP+_ resumes to the ratio=[a+b]/[a+c].The relative sensitivity can take values from zero to infinity. A value below one indicates lower sensitivity on saliva than on NP samples; a value above one indicates that testing on saliva is more sensitive than on the NP swab.

**Table 1.** Contingency table (2×2) for the comparison of saliva and nasopharyngeal (NP) swabs (in absolute numbers).

|  | NP-positive | NP-negative | row total |
| --- | --- | --- | --- |
| saliva-positive | a | b | a+b |
| saliva-negative | c | d | d+d |
| column total | a+c | b+d | a+b+c+d |

A second outcome to be assessed, was the proportion of saliva-positive samples among NP-positive swabs. The inclusion was restricted to patients being enrolled based on a previous NP-positive swab or based on laboratory confirmed COVID-19 disease (a/[a+c], see Table 1). This proportion can take values from zero to 100%.

Meta-analytical pooling using random effect models was performed in Stata (Stat version 16, StataCorp LLC, College Station, TX, USA). The procedure *metan* was applied to pool ratios (i.e. relative sensitivity) and *metaprop* to pool proportions.^20,21^ A continuity correction was applied when a cell of the contingency Table 1 contained a zero value. Heterogeneity among studies was assessed by Cochran’s Q-test (p-value below 0.05 was defined as statistically significant) and by the I^2^ index that describes the proportion of total heterogeneity due to inter-study variation.^22^ Possible influencing covariates were: severity of symptoms, hospitalization status, confirmation status of COVID-19 disease at enrollment and method to collect saliva (swabbing or spitting). Patients who presented respiratory symptoms without dyspnea were categorized as mild whereas patients admitted to an ICU (Intensive Care Unit) with or without the need for oxygen treatment or who died, were categorized in the severe group.

## RESULTS

### Study selection and study characteristics

The last literature search on June 6^th^, 2020 yielded 12 studies of which 9 studies were published in peer-reviewed journals and 3 studies in pre-print journals. In total 1167 patients were included, and 8/12 studies fulfilled the selection criteria for the first outcome.^11,23-29^ Figure 1 summarizes the study selection process. The sample size of these eight included studies varied from 11 to 501 (median=56) and comprised 1070 patients in total. Three hundred and sixteen patients were hospitalized of which 37 were admitted to an ICU (for the study of McCormick-Baw et al.^25^, this information was unknown) and 754 were non-hospitalized. For the second outcome, we pooled the data of eight studies from the first outcome (n=160 patients)^23,24,27,28^ together with the data of four other studies (n=97 patients)^30-33^. Among these patients, 218 were hospitalized and 39 were ambulant. More study characteristics are described in Supplementary Table 1 and Table 2.

**Figure 1.**
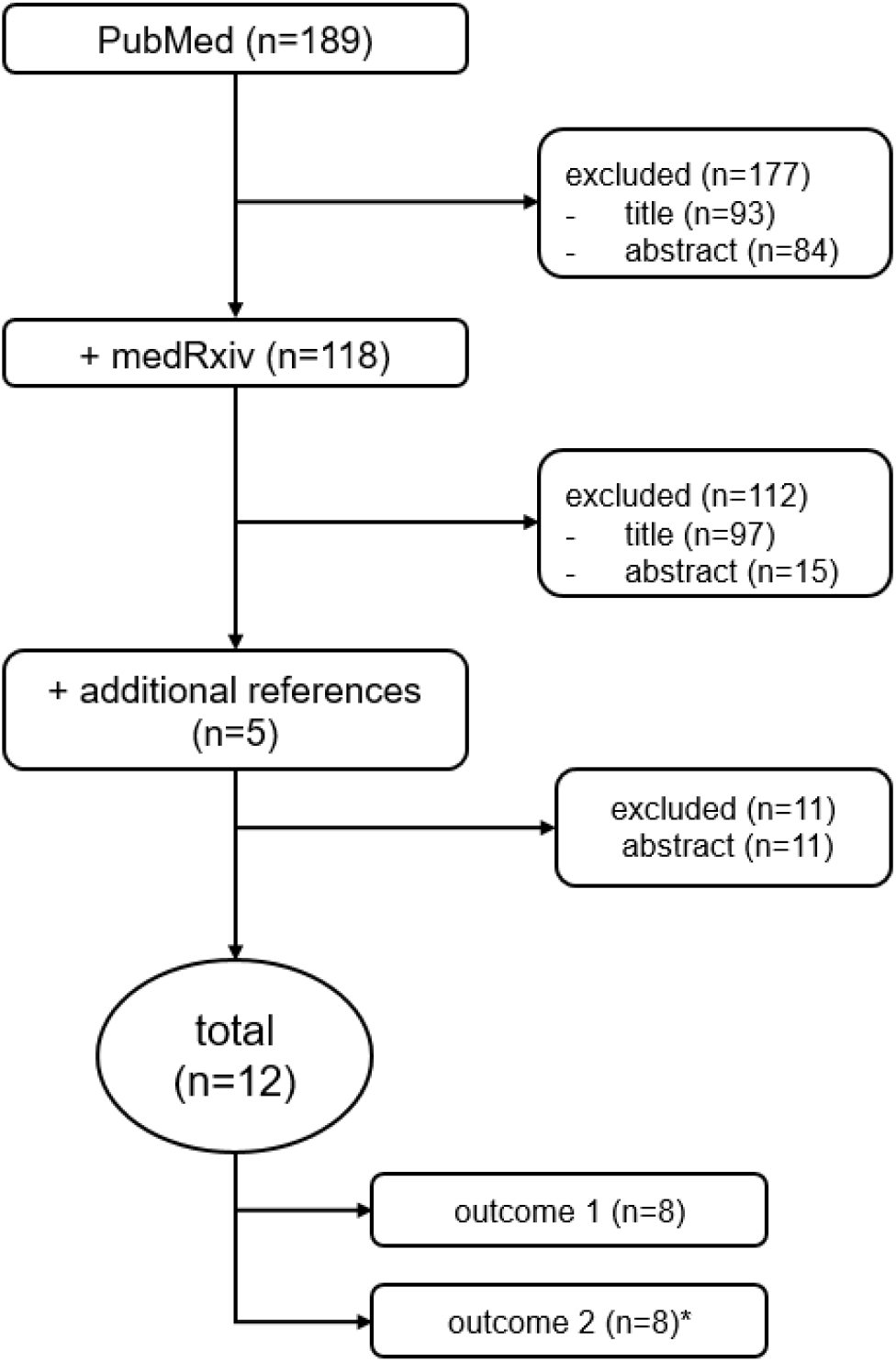
Flow chart: literature search and study inclusions. *The assessment of the second outcome included four studies of the first outcome and additional four other studies.

### Outcome 1: relative sensitivity of SARS-CoV-2 testing on saliva versus nasopharyngeal samples

The sensitivity of SARS-CoV-2 testing was not significantly lower on saliva compared to testing on NP swabs (pooled relative sensitivity was 0.97, 95% CI=0.92-1.02, I^2^=24%) (Figure 2). The relative sensitivity did not differ significantly (p=0.242) by method of saliva collection: 0.98 [95% CI=0.91-1.06] for spitting in vials and 0.94 [95% CI=0.84-1.04] for oral swabbing (Figure 3).

**Figure 2.**
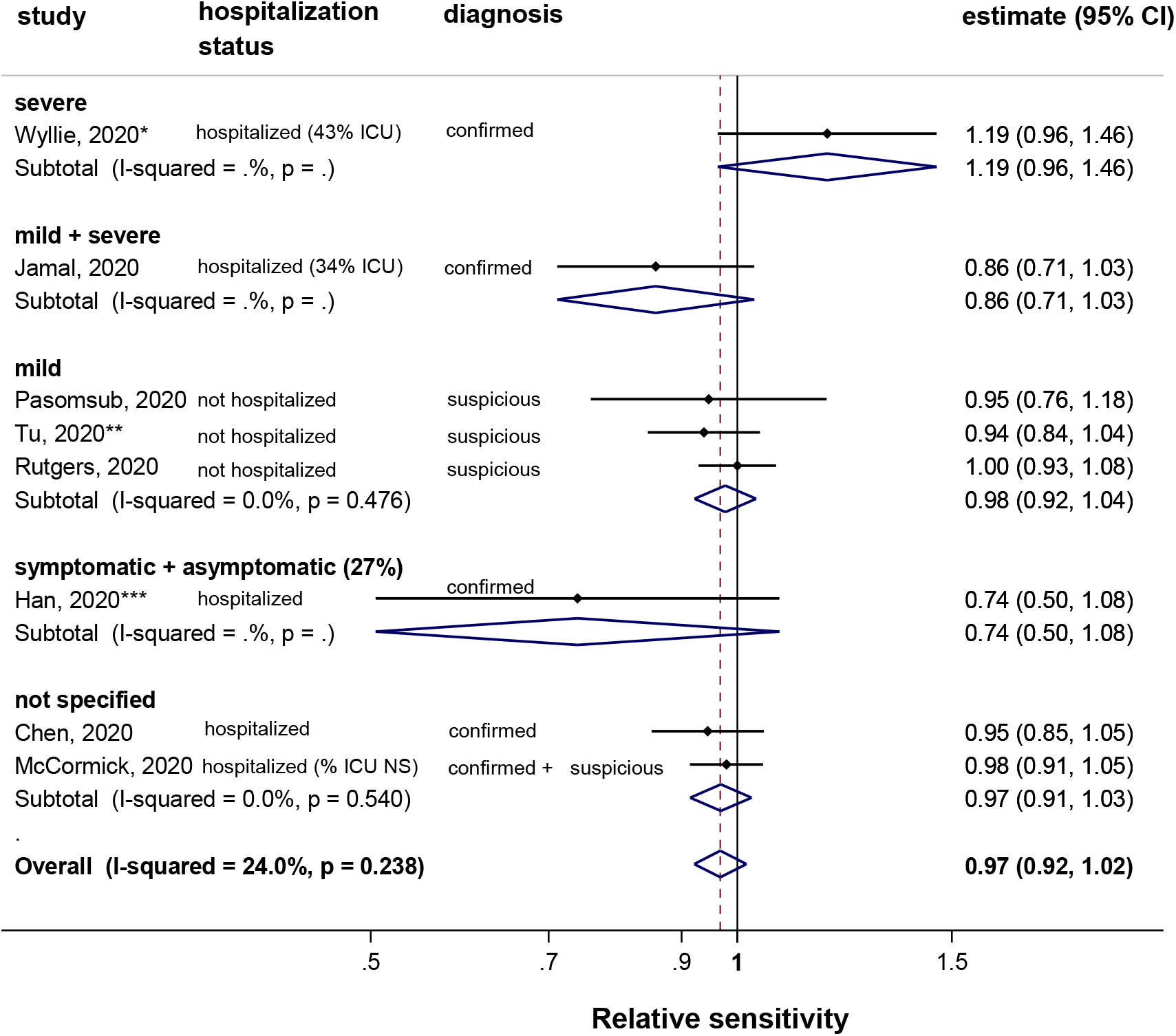
Meta-analysis of the relative sensitivity to detect SARS-CoV-2 RNA in saliva versus nasopharyngeal specimen, stratified by severity of symptoms (in bold). Note: ICU=Intensive Care Unit; NS=not specified. *The proportion of severe patients admitted on ICU was 43% (19/44) for the initial cohort. The patient-paired analysis was performed on a subsample of this cohort (n=38). **Tu et al.^29^ collected a tongue swab. ***We did not subdivide the analysis between asymptomatic (n=3) and symptomatic (n=8) due to the small sample size for each category.

**Figure 3.**
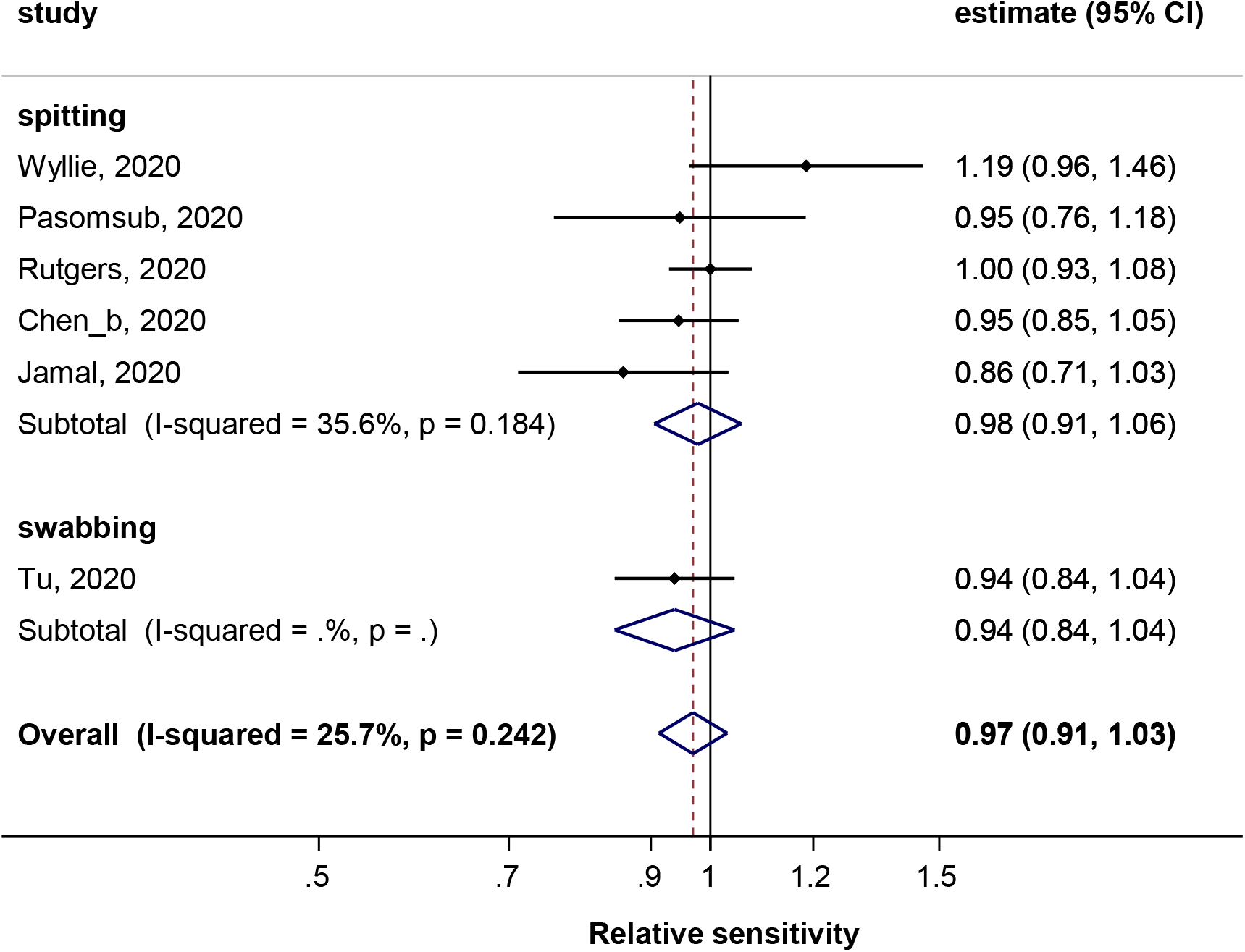
Meta-analysis of the relative sensitivity to detect SARS-CoV-2 RNA in saliva versus nasopharyngeal specimen, stratified by collection method saliva. Note: The studies from Han et al.^23^ and McCormick-Baw et al.^25^ were not plotted because the saliva sampling method was not reported.

The influence of the severity of disease on the relative sensitivity could not be assessed appropriately by lack of strata-specific data. One study found a higher sensitivity in saliva samples than in NP samples, although the 95% CI included unity (1.19, 95% CI=0.96-1.46]. This study included only severe patients of which 43% were admitted to ICU.^27^ The study of Jamal et al.^24^, which included patients with mild or severe symptoms of which 34% was admitted to ICU, found that the sensitivity in saliva samples was 14% lower compared to NP samples, although this was not significant (0.86, 95% CI=0.71 -1.03). In three studies, enrolling non-hospitalized patients with mild symptoms, the pooled relative sensitivity was 0.98 (95% CI=0.92-1.04, p=0.476, I^2^=0%).^11,26,29^ A small study enrolled paired samples from eight symptomatic and three asymptomatic individuals all younger than 17 years, with an overall relative sensitivity of 0.74 [95% CI 0.50-1.08]. The sensitivity on saliva in the symptomatic population was 75% (=6/8) versus 67% (=2/3) in the asymptomatic population.^23^ For two studies, there was no clinical information on disease severity and they had a pooled relative sensitivity of 0.97 (95% CI=0.91-1.03, I^2^=0%).^25,28^

### Outcome 2: proportion of SARS-CoV-2 saliva-positive samples among nasopharyngeal-positive samples

SARS-CoV-2 saliva positivity was reported in four studies that enrolled only NP-positive patients^30-33^. Four other studies from the first outcome that enrolled only COVID-19 confirmed patients^23,24,27,28^ were also included in this assessment. The pooled proportion of saliva-positive samples among NP-positive cases was 86% (95% CI=77-93%, Figure 4). There was substantial heterogeneity among the studies (I^2^=66%, p<0.05). The highest proportion was found in the group with exclusively severe disease (93%, 95% CI=85-98%). A lower proportion was found in the mixed group with mild and severe disease (73%, 95% CI=63-83%) which was similar to the proportion of the other mixed group with symptomatic and asymptomatic cases (73%, 95% CI=43-90%). The results of To et al.^31^ could be further separated between mild and severe disease which resulted in a slightly higher proportion of saliva-positive samples among severe cases (88%, 95% CI=53-98%) compared to mild cases (85%, 95% CI=58-96%, p=0.85).

**Figure 4.**
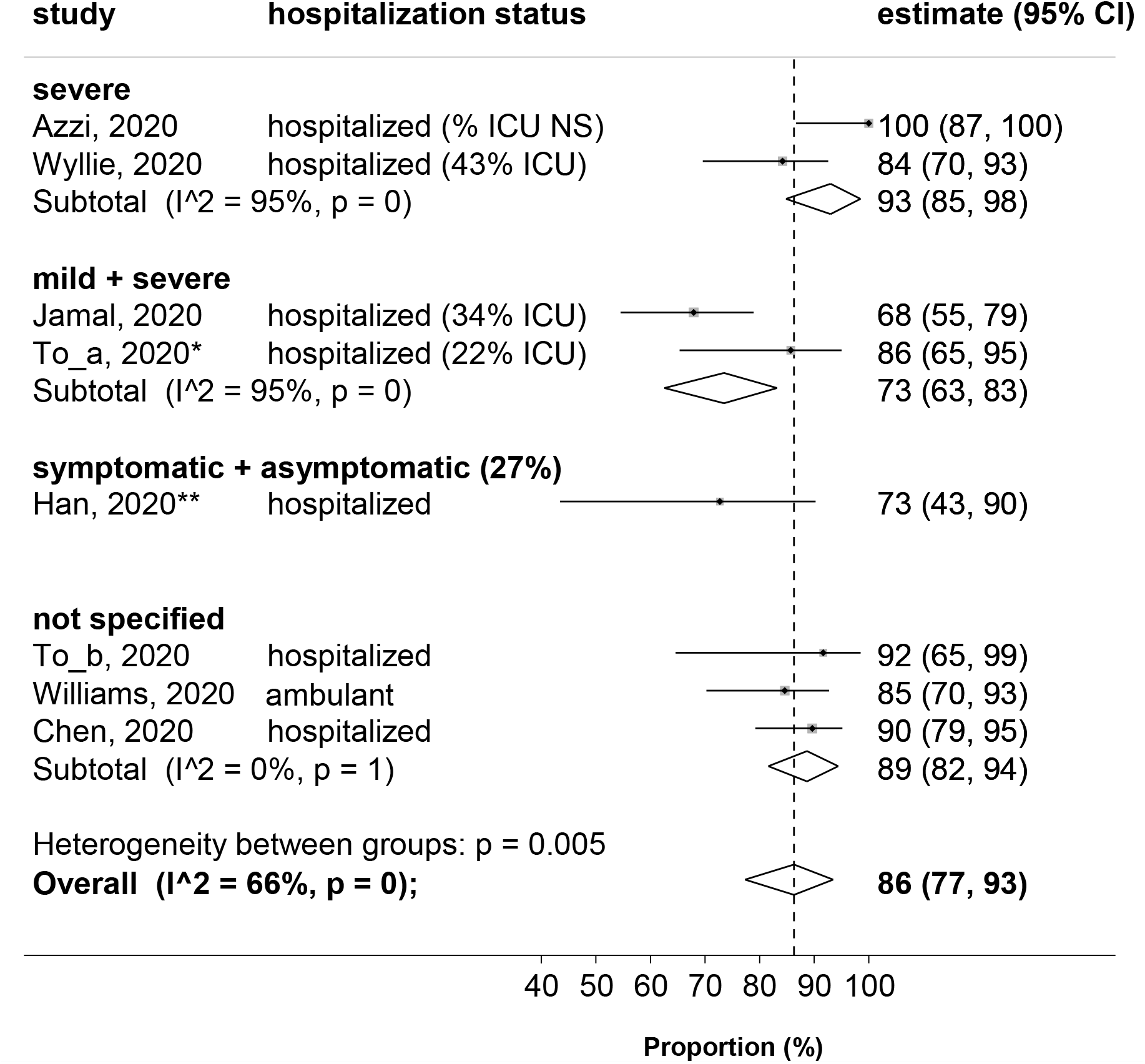
Proportion (%) of saliva-positive samples among nasopharyngeal-positive samples. Note: ICU=Intensive Care Unit; NS: not specified. The COVID-19 confirmation was based on a positive NP or oropharyngeal swab for the studies of Han et al.^23^, Wyllie et al.^27^ and Chen et al.^28^ For the study of To_b et al.^30^ this was based on a positive NP or sputum sample. For the other studies^33,24,31,32^ this was based on a NP-positive sample. *The ICU-percentage was based on a study population of 23, whereas the percentage of mild cases and proportion was based on a study population of 21, because there was no study data for two patients who died during the study.^31^ Twelve patients were also included in To_b et al.^30^ **We did not subdivide the analysis between asymptomatic (n=3) and symptomatic (n=8) due to the small sample size for each category.

## DISCUSSION

Our meta-analysis did not reveal a significant difference in virological sensitivity of SARS-CoV-2 RT-qPCR testing on saliva compared to NP swab transport medium. On average 14% of patients who were positive for SARS-CoV-2 using NP swabs tested negative when a saliva sampling method was used.

### Limitations of the systematic review

The retrieved literature focuses mainly on the virological sensitivity to detect a SARS-CoV-2 infection in different specimens. Additionally, there may have been implicit differences in method of collection, laboratories and equipment used that could not be investigated thoroughly due to limited information available. Besides, specificity to exclude an infection was rarely reported. Specificity is also an important parameter to consider particularly in situations where the pandemic is less prevalent and testing is used primarily for screening and contact-tracing purposes.

The estimation of the virological accuracy of SARS-CoV-2 testing on saliva is only reliable when the RT-qPCR assays themselves are properly validated. This will further be evaluated in the VALCOR ("VALidation of SARS-CORona Virus-2 assays”) study which is currently ongoing in Belgium.^34^ The included studies comprised a heterogeneous case mix of subjects (from asymptomatic to patients with severe symptoms admitted to ICU). However, the sources of heterogeneity could not be assessed by lack of separate test results stratified by covariates. Furthermore, we encountered a lack of data for the second outcome with regard to the specifications of the RT-qPCR assays (i.e. manufacturer’s name and target genes) that have been used for the confirmation of on NP samples. Therefore, it might be possible that the RT-qPCR that has been used a priori on NP samples was different from the assay applied on saliva samples and hence causes biases.

Certain studies^23,32^ showed that the viral load was lower in saliva than on NP, whereas Wyllie et al.^27^ found a higher viral load in saliva. In order to extend our findings with information on the timing of collection, we propose to investigate the difference in viral load and duration of viral shedding between saliva and NP swabs when more studies with larger sample sizes are published.

Our study results must be interpreted with caution. In particular, we cannot assure that SARS-CoV-2 testing on saliva is sufficiently sensitive among recovering patients or among asymptomatic individuals.

Inherent to meta-analyses based on pooling of aggregated information published in papers, the role of covariates often cannot be evaluated precisely by the lack of homogenously stratified data.^35^ It might be worthwhile to perform individual-patient data (IPD) meta-analyses. However, obtaining IPD is difficult and biases may occur when only a limited number of authors consent to sharing data.

More studies with alternative procedures for sample collection and specimen handling, spanning a wide range of patient groups, including asymptomatic individuals, preferentially with similar protocols are needed in order to facilitate future pooling and enhance our understanding of using different samples for SARS-CoV-2 testing. The number of studies contributing to our meta-analyses is small, precluding formal statistical assessment of publication bias. However, given the very high desirability to find quicker, safer and more comfortable collection procedures, publication bias can be presumed. In line with this, preliminary results in the Belgian federal task force led study on saliva (n=2000) indicates that viral load is lower in saliva than in NP swab transport medium (using Cq as a proxy for viral load), and that good correspondence between saliva and NP swab is only observed for medium to high viral loads in NP (manuscript in preparation).

## CONCLUSION

In conclusion, we found that SARS-CoV-2 testing by RT-qPCR on saliva appears to be as sensitive as NP swabs. However, the quality of included studies often was low and lacked stratified data, which impeded assessment of the sources of heterogeneity. Therefore, results from our study should be interpreted with caution. More information is needed before saliva can be recommended as a valid diagnostic alternative for NP samples.

## Data Availability

Data is available upon request.

## ACKNOWLEDGEMENTS

EP, SKDAS and MA would like to acknowledge the VALCOR project^34^ funded by emergency funding from the Federal Belgian Government. Biogazelle is part of the SARS-CoV-2 testing task force, set up upon request of the Belgian government to increase testing capacity during the COVID-19 pandemic in Belgium.

## SUPPLEMENTARY MATERIALS

Supplementary Table 1 and Table 2.

